# Youth in crisis: comparative geographies of suicide between Argentina, Chile, Spain and Uruguay

**DOI:** 10.64898/2026.02.06.26345682

**Authors:** Carlos M. Leveau, Pablo Hein Picó, Ana Santurtún

## Abstract

**Introduction:** National trends in youth suicide risk may mask significant regional variations within a country. This article attempts to account for spatio-temporal trends through a comparative analysis across South America and Europe. This paper analyzes the spatiotemporal patterns in suicide mortality among young people (10-29 years) in Argentina, Chile, Spain, and Uruguay during the period 1997-2021.

**Methods:** Official data from vital statistics and population censuses of the four countries were analyzed. Spatiotemporal clusters were detected using Poisson-based scan statistics. Sociodemographic characteristics of high-and low-mortality clusters were compared with the rest of each country using Kruskal–Wallis and Wilcoxon tests.

**Results:** With the exception of Chile, each country showed the emergence of spatiotemporal suicide clusters extending through 2021. Indicators of social fragmentation and lower socioeconomic status were most consistently associated with the formation of high-risk youth suicide clusters.

**Conclusion:** Recent national increases in youth suicide rates appear to be concentrated in specific sub-national regions, underscoring the need to target resources toward improving living conditions and mental healthcare access for young people in these areas.

## Introduction

Suicide is a complex phenomenon shaped by the interplay of psychiatric, psychological, demographic, socioeconomic, and cultural factors (Monk, 1987; Wray et al., 2011). In contrast to the global decline in suicide rates, the Americas is the only region experiencing a sustained increase (Ilic and Ilic, 2022). Suicide is the third leading cause of death among people aged 15–29 globally—the second leading cause for women and the third for men in this age group (Organization, 2025). These global trends, however, are characterized by significant geographical and temporal variation. While Europe has experienced a decline in youth suicide rates over the past three decades, the Americas have seen a steady increase in recent years (Bertuccio et al., 2024).

These global variations highlight the critical influence of contextual factors in shaping recent trends in youth suicide. While individual risk factors are essential, the geographical variation in youth suicide rates may be largely explained by regional differences in socio-demographic, economic, and cultural contexts (Stack, 2021). Following Durkheim (Durkheim, 2002), social fragmentation—marked by weakened community ties, secularization, evolving family structures (such as divorce, single-parent households, and living alone), greater residential mobility, and the decline of traditional social institutions—tends to reinforce individualism and is associated with rising suicide rates among youth (Ghadipasha et al., 2024; Stack, 2021; Stockard and O’Brien, 2002; Trovato, 1992). Although these social fragmentation processes are often more intense in urbanized settings, rural areas exhibit higher rates of youth suicide (Ghadipasha et al., 2024). Socio-spatial isolation and poor access to health services –particularly mental health services– are characteristic factors of rural areas associated with a higher risk of youth suicide. Rural areas also present distinct cultural factors –including stoicism and self-efficacy that discourage help-seeking for mental distress– which can make suicide appear as a viable solution to personal crises (Judd et al., 2006). Areas of low socioeconomic status show elevated youth suicide rates (Ghadipasha et al., 2024; Gould et al., 2003), where populations face not only material hardship but also higher exposure to multiple forms of violence, territorial stigma, job insecurity, and inadequate access to mental health care.

These sociodemographic factors are not geographically uniform. Therefore, national trends in youth suicide risk may mask significant regional variations within a country. This article attempts to account for these spatio-temporal trends through a comparative analysis of four countries – Argentina, Chile, Spain and Uruguay. In addition, the socio-demographic characteristics of the areas associated with the observed spatiotemporal trends will be examined. While a limited number of national studies exist (Alarcão et al., 2019; Leveau and Alazraqui, 2020; Núñez-González et al., 2018), there is a gap in the literature regarding recent spatiotemporal trends in youth suicide across South America and Spain.

While Spain and the Southern Cone nations share cultural similarities—Spanish language and a predominant Catholic heritage—there are variations in the degree of secularization among the South American countries in the region. While 17% of youth in Argentina report no religious affiliation (Latinobarómetro, 2023a), this rises to 49% in Chile (Latinobarómetro, 2023b) and 66% in Uruguay (Corporación Latinobarómetro, 2024). The four countries also differ in other socio-cultural and economic dimensions. Unlike Spain, Argentina, Chile, and Uruguay have populations of Indigenous descent or affiliation, though their demographic weight varies across the three nations. Uruguay reports the lowest share of its population identifying as Indigenous or of Indigenous descent (under 3%), followed by Argentina (10%) and Chile (13%) (Sans, 2022). In terms of socio-economic development, Spain maintains a more comprehensive welfare state—especially in health and education—and exhibits lower social inequality compared to Argentina, Chile, and Uruguay. Spain’s GDP per capita (US$35,327) exceeds that of the three South American countries, which range from a low of US$13,970 (Argentina) to a high of US$23,907 (Uruguay) (World Bank, 2025). At the same time, income inequality is lower in Spain (Gini index: 33.4) than in the three South American nations, where it ranges from 40 to 43 (World Bank, 2025). These parallels and contrasts in both cultural characteristics and socio-economic conditions present a valuable case study for examining the contemporary geography of youth suicide across varied national contexts.

Therefore, the overall objective is to analyze, from a spatio-temporal perspective, suicide mortality among young people in Argentina, Chile, Spain, and Uruguay during the period 1997-2021. The specific objectives are: (a) to discover the formation of spatio-temporal clusters of suicides among young people; and (b) to examine the role of sociodemographic factors on suicide mortality among young people.

## Methods

Data from official national sources—the ministries of health and national statistical institutes of Argentina, Chile, Spain, and Uruguay—were used for this study (Instituto Nacional de Estadísticas - Chile, 2024; Ministerio de Salud - Argentina, 2026; Ministerio de Salud Pública - Uruguay, 2026). The databases included records of suicide mortality (ICD-10 codes X60.0–X84.9) by area of residence, sex, and age. Data were aggregated at the province level for Argentina, Chile, and Spain, and at the department level for Uruguay, covering the period 1997–2021. Due to administrative boundary changes in Chile over the study period, a harmonized map was created to ensure temporally consistent spatial units (Table S1 in the Supplementary Material). Youth suicide was defined as occurring between ages 10 and 29. In Argentina, persistent underreporting of violent deaths suggests that some deaths classified as injuries of undetermined intent (DUI) likely correspond to misclassified suicides (Santoro, 2020). Among DUIs, deaths due to undetermined injury by hanging, strangulation, or suffocation (Y20) are most likely to represent misclassified suicides (25). Therefore, these cases were combined with confirmed suicides for analysis (Table S2, Supplementary Material). In Chile, underreporting of suicide was also apparent, particularly in the Santiago Metropolitan Region during 1997–2000. A similar adjustment was applied, adding deaths coded as Y20 to the suicide counts (Table S2, Supplementary Material).

To calculate suicide rates, census population data (from rounds near 2000, 2010, and 2020) and annual intercensal population estimates were used (Instituto Nacional de Estadística, 2025a; Instituto Nacional de Estadística - Uruguay, 2025; Instituto Nacional de Estadística y Censos de la República Argentina, 2025a, 2025b; Instituto Nacional de Estadísticas - Chile, 2024; Instituto Nacional de Estadísticas de Chile, 2025). In Argentina, official annual population projections are only available from 2010 onward. Therefore, linear population estimates for the 1997–2009 period were derived using census data from 1991, 2001, and 2010. In Chile, official population projections from the National Institute of Statistics were used for 2002–2020. For 1997–2001, linear estimates were derived from the 1992 census. Since official projections ended in 2020, the 2020 estimate was applied to 2021 as well. In Spain, official population projections from the National Institute of Statistics covered 1998–2021; the 1998 figure was therefore applied to 1997. For Uruguay, intercensal estimates were used for 1997– 2011 and official projections for 2012–2021 (Instituto Nacional de Estadística - Uruguay, 2025).

Spatiotemporal scan statistics (Kulldorff et al., 1998) were applied using moving cylindrical windows with a geographic base and a temporal height, which scanned the entire study area. Under the discrete Poisson model—which assumes a uniform spatial and temporal distribution of suicide risk—observed cases were compared against expected cases, adjusted for population. The alternative hypothesis identifies cylinders (spatiotemporal windows) with significantly elevated or reduced risk (p < 0.05, assessed with 9999 Monte Carlo simulations). The most likely clusters—those with the greatest excess of cases—were detected using SaTScan software version 9.1.1 (Kulldorff, Harvard Medical School).

Finally, sociodemographic characteristics were compared among three groups: (1) areas within high-mortality clusters, (2) areas within low-mortality clusters, and (3) the remaining territory in each country, using Kruskal-Wallis tests. In cases where only high-mortality clusters were identified, two-sample Wilcoxon tests were used to compare them against the rest of the territory. When statistically significant differences were found, medians and interquartile ranges were reported. Four harmonized socio-demographic indicators, available for all four countries, were compared: population density (inhabitants per km^2^) as a measure of population concentration; the percentage of adults with completed university or tertiary education as an indicator of area-level socioeconomic status; the percentage of adults who are single, divorced, or widowed (i.e., unmarried or without a partner); and the percentage of single-person households. The latter two variables served as proxies for the degree of social fragmentation in each area. All variables were sourced from national population censuses (Instituto Nacional de Estadistica, 2025; Instituto Nacional de Estadística de Uruguay, 2024; Instituto Nacional de Estadística y Censos, 2026; Instituto Nacional de Estadísticas - Chile, 2025) or national surveys (“Encuesta Continua de Hogares (2020). Número de hogares por provincias según tamaño de hogar y número de habitaciones de la vivienda,” 2025; Instituto Nacional de Estadística, 2025b). Detailed information on data sources is provided in Table S3 (Supplementary Material). Statistical analyses (Kruskal–Wallis and Wilcoxon tests) were conducted using Stata/SE 13.1, while spatiotemporal cluster mapping was performed in QGIS Desktop 3.16.7.

## Results

During 1997–2021, there were 48,530 suicides among men and 12,645 among women across the four countries. Argentina recorded the highest absolute numbers for both sexes (men: 26,712; women: 7,157), while Uruguay had the lowest (men: 2,786; women: 612). In terms of average annual suicide rates per 100,000 population, Uruguay showed the highest rates for both women (4.8) and men (21.4), followed by Argentina (women: 4.3; men: 16.0), Chile (women: 3.9; men: 15.3), and Spain, which had the lowest rates (women: 1.5; men: 5.5).

Figure 1 shows the annual variations in youth suicide rates across the four countries. In Argentina, suicide rates for both women and men peaked in 2002. After several years of relative stability, rates in both sexes began to fluctuate and rise toward the end of the study period. In Chile, suicide rates reached their highest levels during 2007–2010. Following this peak, rates for both sexes stabilized, though for men they remained lower than the levels observed in 1997–2006. In Spain, the suicide rate among young women reached its lowest point in 2010–2011 before returning to earlier levels. Among young men, the rate also reached a low point in 2010, but it appears to have followed a general downward trend from 1997 to 2021. In Uruguay, suicide rates among both young men and women displayed an overall upward trajectory. The rate for men peaked in 2002, showing a similar temporal pattern observed in Argentina.

**Figure 1.**
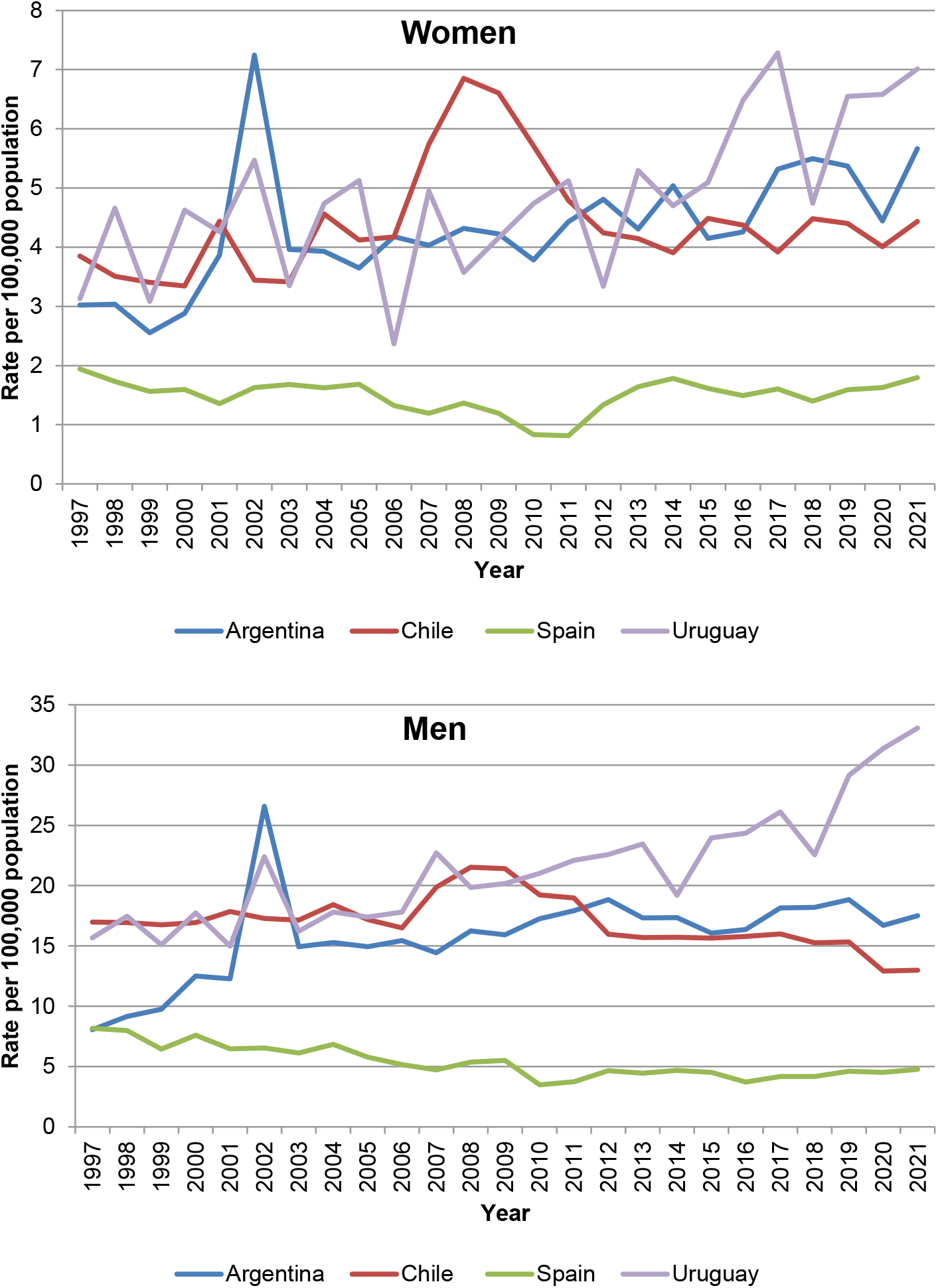
Temporal variations in suicide rates among young people (10-29 years) in women and men. Argentina, Chile, Spain and Uruguay, 1997-2021.

Spatiotemporal scan analysis detected significant clusters of elevated and reduced youth suicide risk among both women and men in all four countries. Figure 2 presents these clusters for Argentina. For both sexes, northern Argentina emerged as a high-risk region during the latter part of the study period (1997–2021). For women, the COVID-19 pandemic appeared to interrupt the formation of a significant cluster of high youth suicide risk in the northwestern provinces (Figure 2, cluster 1).

**Figure 2.**
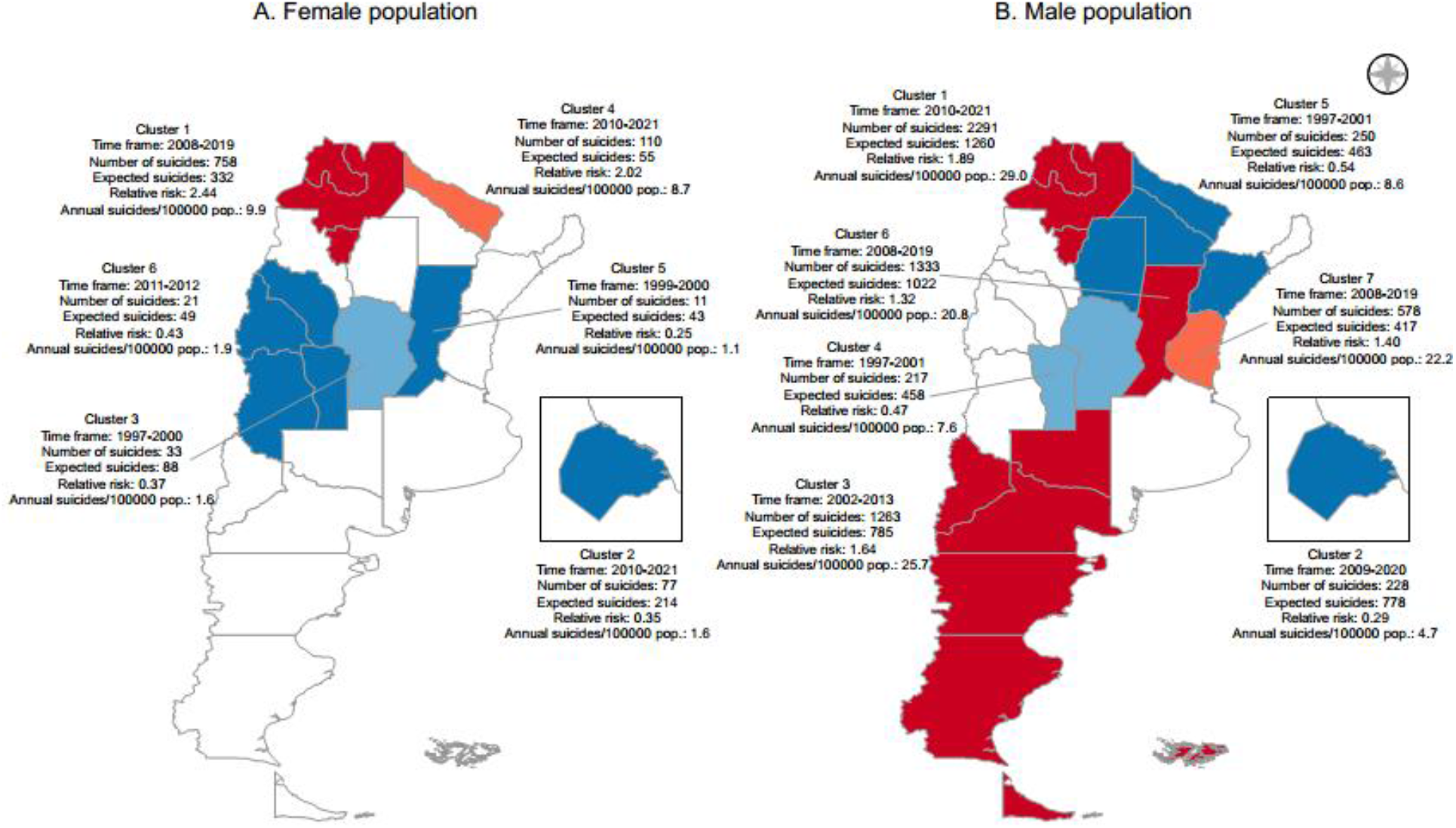
Spatio-temporal clusters of high (red tones) and low (blue tones) risk of youth suicide in Argentina, 1997-2021.

Figure 3 shows the spatiotemporal concentrations of high and low risk of youth suicide in Chile. While clusters of high suicide risk emerged throughout the country in both sexes, no spatial concentrations of high suicide risk emerged in the later years of the study period.

**Figure 3.**
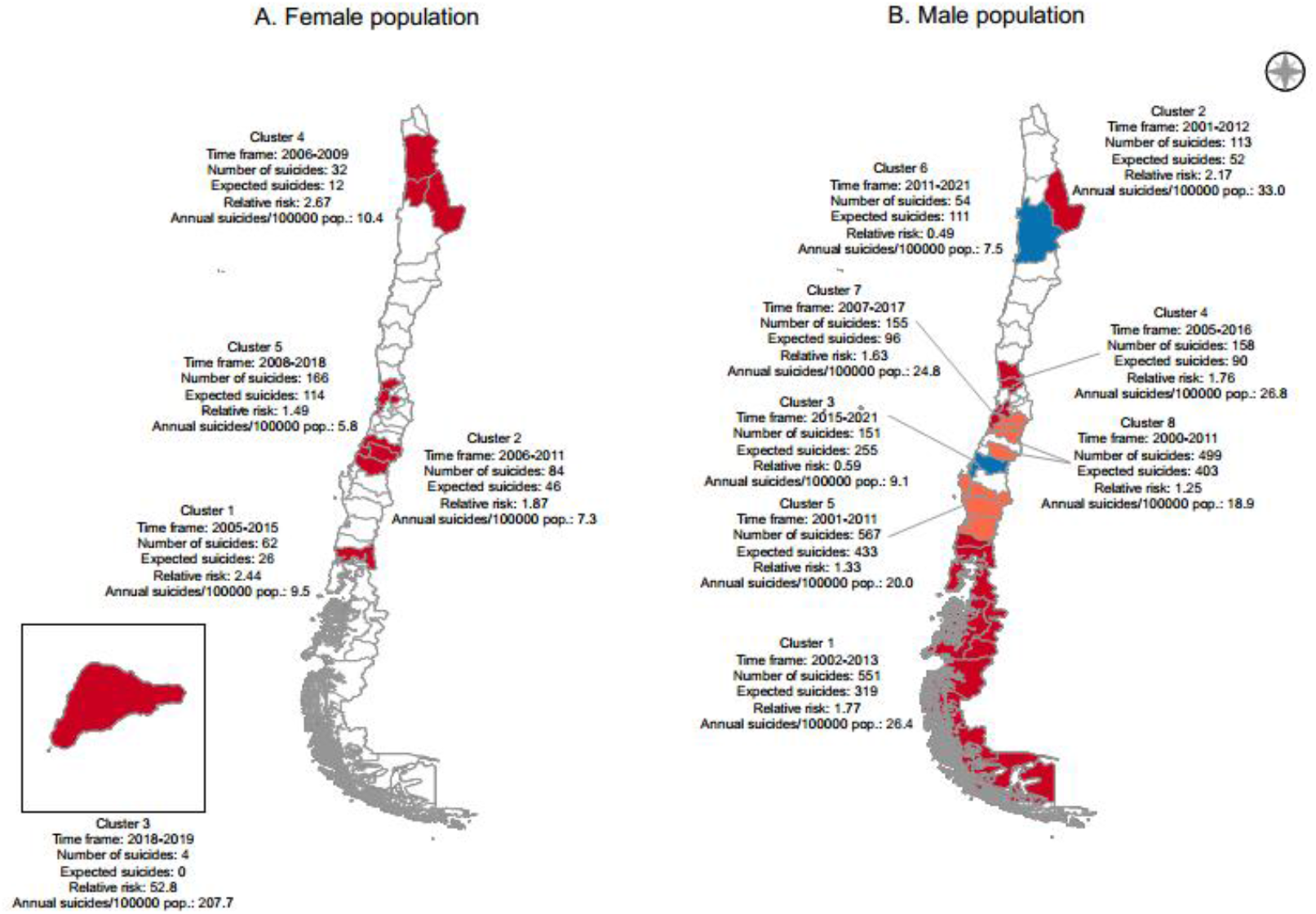
Spatio-temporal clusters of high (red tones) and low (blue tones) risk of youth suicide in Chile, 1997-2021.

Figure 4 shows the spatiotemporal concentrations of high and low risk of youth suicide in Spain. Compared to Argentina and Chile, Spain showed greater spatiotemporal differences between women and men. For women, a single high-risk cluster emerged in the northwest during 2012–2021. For men, five high-mortality clusters were identified, located in both northern and southern regions. Four of these male clusters spanned—at minimum—the period 1997–2005.

**Figure 4.**
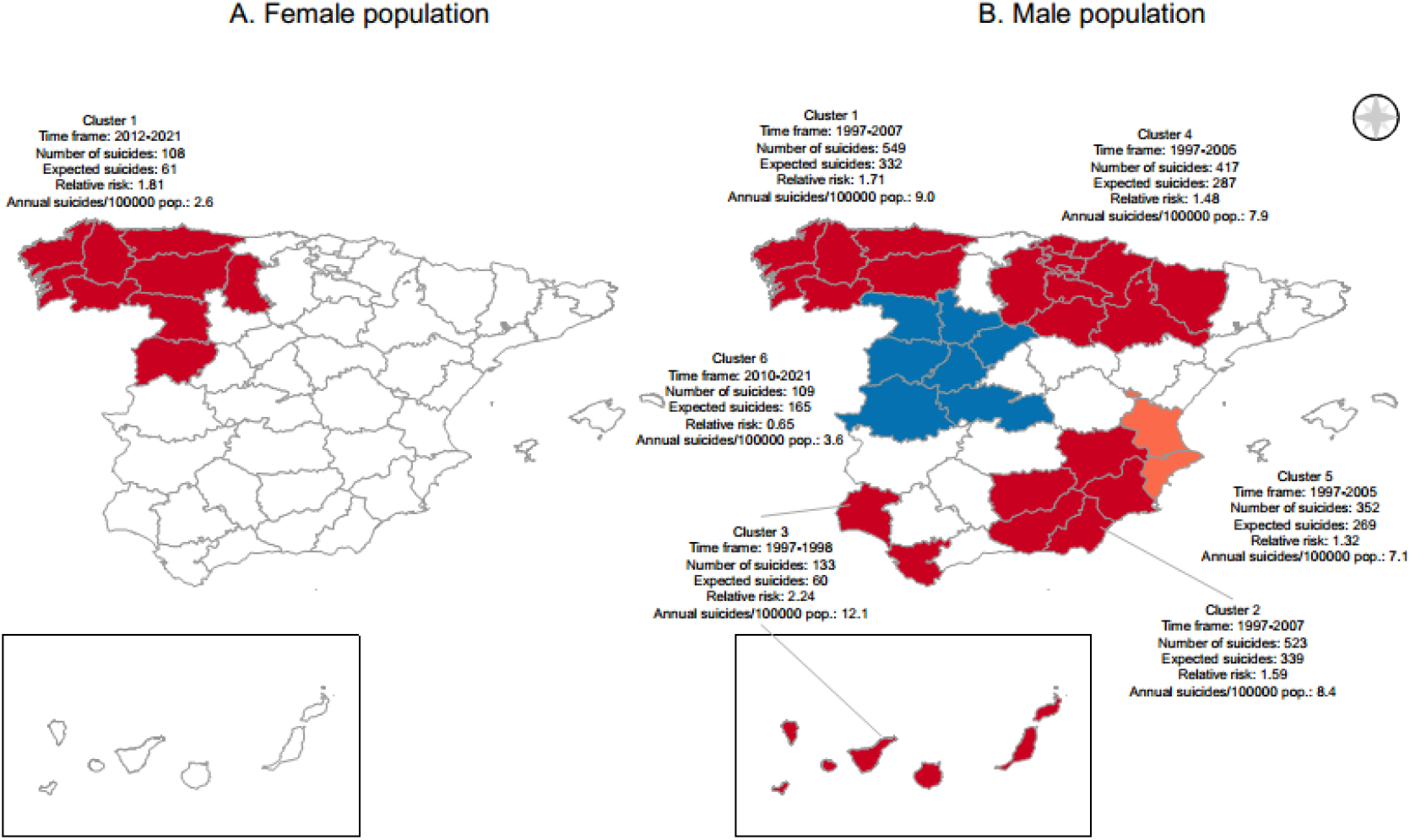
Spatio-temporal clusters of high (red tones) and low (blue tones) risk of youth suicide in Spain, 1997-2021.

Finally, in Uruguay, three high-risk clusters of youth suicide emerged among males, all during the latter years of the study period (1997–2021), located in both eastern and western regions and excluding the most urbanized departments (Montevideo, Canelones, and Maldonado) (Figure 5). For females, two high-risk clusters emerged in western Uruguay, though they were more limited in both geographic extent and duration compared to the male clusters (Figure 5).

**Figure 5.**
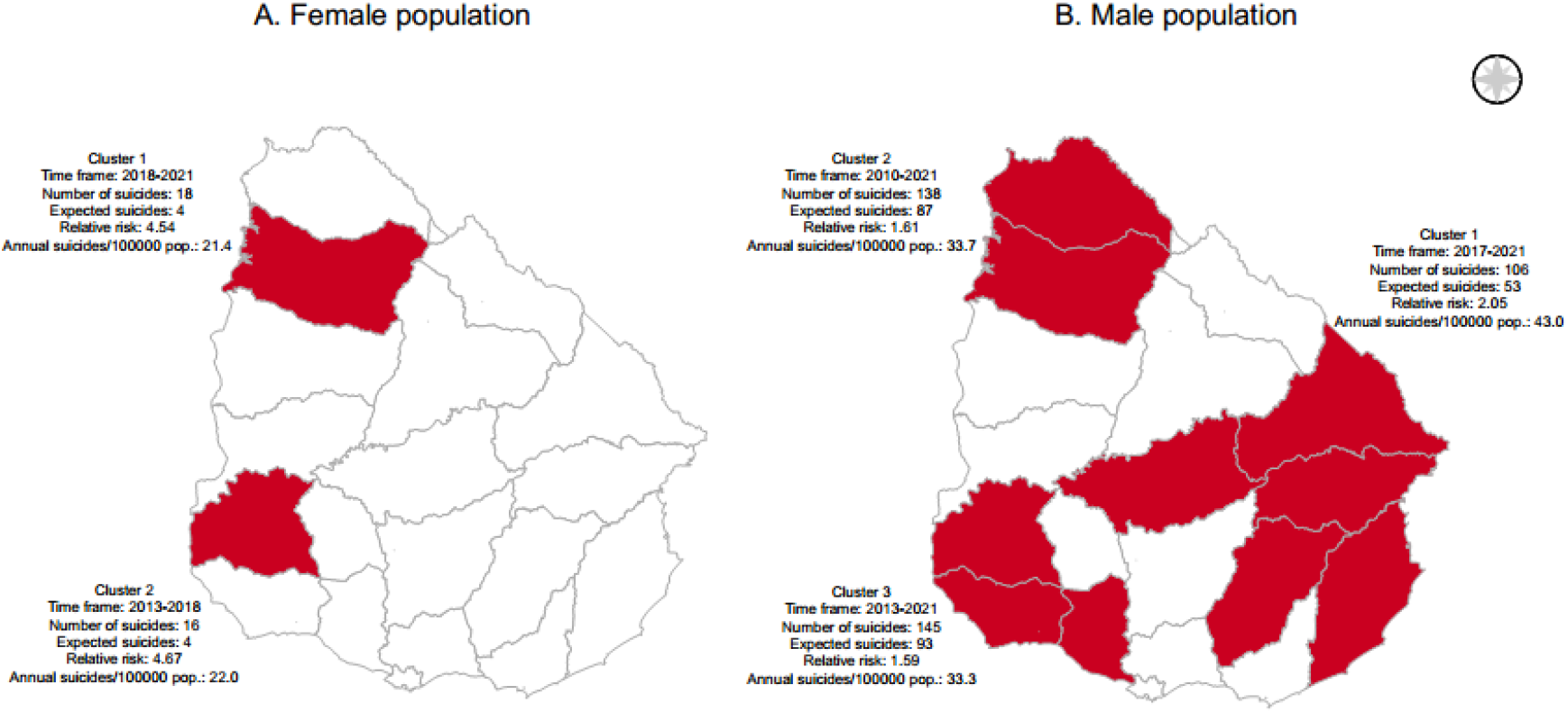
Spatio-temporal clusters of high (red tones) and low (blue tones) risk of youth suicide in Uruguay, 1997-2021.

In Argentina, statistically significant differences in the proportion of unmarried population and in educational attainment were observed among three types of areas for the female population: high-risk clusters, low-risk clusters, and the rest of the territory (Table 1). The median percentage of unmarried adults was highest in high-risk clusters (70.7, IQR: 66.5–72.9), followed by the rest of the country (66.9, IQR: 65.1– 67.9), and lowest in low-risk clusters (61.7, IQR: 60.3–65.3). Educational level showed the opposite pattern: lowest in high-risk clusters (12.8%, IQR: 12.1–12.9), intermediate in the rest of the country (14.5%, IQR: 13.3–15.3), and highest in low-risk clusters (16.0%, IQR: 13.7–17.9).

**Table 1.** Kruskal-Wallis (H) and Wilcoxon (U) tests, Argentina, Chile, Spain and Uruguay, 1997-2021.

| Sex | Variables | Argentina | Chile | Spain | Uruguay* |
| --- | --- | --- | --- | --- | --- |
| Women | Population density | H=3.468; p=0.177 | U=-1.294; p=0.1957 | U=1.073; p=0.2832 | - |
|  | Unmarried population | H=6.986; p=0.0304 | U=0.343; p=0.7317 | U=2.285; p=0.0223 | - |
|  | Single-person households | H=5.418; p=0.0666 | U=-0.542; p=0.5879 | U=-2.689; p=0.0072 | - |
|  | Educational level | H=7.640; p=0.0219 | U=-0.431; p=0.6663 | U=-0.164; p=0.8696 | - |
| Men | Population density | H=1.785; p=0.410 | U=1.418; p=0.1563 | H=6.618; p=0.0366 | U=0.735; p=0.4624 |
|  | Unmarried population | H=1.843; p=0.398 | U=1.340; p=0.1803 | H=7.199; p=0.0273 | U=-0.653; p=0.5136 |
|  | Single-person households | H=2.959; p=0.2278 | U=-1.437; p=0.1507 | H=5.201; p=0.0742 | U=-0.653; p=0.5136 |
|  | Educational level | H=1.041; p=0.5943 | U=3.010; p=0.0026 | H=0.811; p=0.6666 | U=1.878; p=0.0604 |

Notes: Argentina, women: areas in high-risk clusters (HR)=4, areas in low-risk clusters (LR)=7, areas in the rest of the country (RC)=13; men: areas HR=11, LR=7, RC=6. Chile, women: areas HR=13, RC=37; men: areas HR=26, LR(n=2)+RC(n=22)=24. Spain, women: areas HR=9, RC=41; men: areas HR=26, LR=7, RC=17. Uruguay, women: areas HR=2, RC=17; men: areas HR=9, RC=10. \*Analysis not performed on women due to only two departments having clusters.

In Chile, the only statistically significant difference was found for educational level among males (Table 1). The median proportion with university or tertiary education was lower in high-risk clusters (21.0%, IQR: 17.0–23.5) than in the combined group of low-risk clusters and the rest of the country (26.6%, IQR: 21.2–31.2). In Spain, among the female population, statistically significant differences were found for the percentage of unmarried individuals and the percentage of single-person households. The median proportion of unmarried adults was lower in high-risk clusters (54.5%, IQR: 53.3–54.7) than in the rest of the country (56.4%, IQR: 54.8–57.7). Conversely, the median percentage of single-person households was higher in high-risk clusters (29.9%, IQR: 27.8–30.6) compared to the rest of the country (26.2%, IQR: 24.6–27.7). For men, significant differences were observed in population density and, as with women, in the percentage of unmarried individuals (Table 1). Median population density was lower in low-risk clusters (22.2 persons/km^2^, IQR: 19.7–46.2) than in both high-risk clusters (78.2 persons/km^2^, IQR: 42.0–210.4) and the rest of the country (88.6 persons/km^2^, IQR: 24.9–138.9). A similar pattern was seen for the proportion of unmarried adults, which was lower in low-risk clusters (54.4%, IQR: 53.7–54.8) compared to high-risk clusters (56.3%, IQR: 54.6–57.3) and the rest of the country (56.8%, IQR: 54.7–58.5). Finally, in Uruguay there were no statistically significant differences in men. For females, statistical testing could not be performed because the two high-risk clusters identified each consisted of only one department (Table 1).

## Discussion

This study showed spatiotemporal variations in youth suicide across Argentina, Chile, Uruguay (Southern Cone of South America), and Spain (Europe) between 1997 and 2021. Despite differing national-level trends, all countries except Chile showed localized areas with persistently high suicide rates that persisted through 2021. Among the three area-level sociodemographic factors examined, those reflecting social fragmentation and socioeconomic status were most consistently associated with the emergence of high-risk spatiotemporal clusters of youth suicide. This study has three principal strengths: 1) its extensive geographic (four countries with a combined population of 119 million) and temporal (25-year) coverage of suicide data; 2) to our knowledge, it is the first comparative spatial analysis of suicide across four countries on two continents; and 3) it examines the influence of area-level risk factors that have been insufficiently studied in youth populations.

An emergence of high-risk clusters of youth suicide in low socioeconomic areas was to be expected (Ghadipasha et al., 2024; Gould et al., 2003). However, this association was not observed in all four countries. In Argentina and Chile, clusters of high-risk suicide among young women and men, respectively, tended to be located in areas of lower socioeconomic status compared to the rest of the country. Unlike Spain, which showed no association between socioeconomic status and the emergence of high-risk suicide clusters, both countries have indigenous populations, which are primarily located in provinces with lower economic development. In Chile, cluster 3 of young female suicides was located on Isla de Pascua (Easter Island), with a population of 7750 inhabitants, of whom 54% belong to indigenous peoples (Chile, 2023). The youth of Easter Island (Rapa Nui) are experiencing a loss of traditional cultural values and a shift toward modern values prioritizing economic success and individualism, a transition that has coincided with rising rates of alcoholism (Pérez, 2004). In Argentina, journalistic reports describe similar problems –drug addiction and violence– in provinces of northwestern Argentina (Jujuy, 2012; López, 2024). However, in both Argentina and Chile, we are unaware of any studies analyzing suicide rates specific to Indigenous populations. In Uruguay, the Wilcoxon test for males resulted in a p-value of 0.06, indicating a trend toward lower educational attainment in high-risk youth suicide clusters. The median percentage of the population with university or tertiary education was lower in these clusters (9.6%, IQR = 7.8–13.4) than in the rest of the country (12.5%, IQR = 8.9–26.5). Therefore, the association between suicide and socioeconomic disadvantage appears strongest in the two countries with the lowest income levels and greatest inequality—Argentina and Chile—and is consequently most pronounced in the poorest regions across the four nations.

Several studies have reported an increase in the youth suicide rate during economic crises (Baeza et al., 2022; Chang et al., 2013; Harper et al., 2015). While, with the exception of Spain, our results showed that temporal trends in youth suicide appeared to be associated with economic crises, no high-risk clusters of suicide emerged in response to these crises. In Argentina, every year of the 1998–2002 period was marked by economic recession, representing the most severe socio-economic and institutional crisis of the entire study period. At the peak of this crisis—2002—there was a spike in youth suicides, representing the highest rate in the entire 1997–2021 period. However, no spatio-temporal clusters of high youth suicide mortality emerged during these years. This suggests that the 2002 crisis was associated with a broadly distributed, nationwide increase in suicide risk across Argentine provinces rather than with localized geographic patterns. Chile, with a more stable economy, experienced two years of economic recession (1999 and 2009), the latter intensified by the 2008 global financial crisis. Nationally, this crisis affected males aged 15-24 (Baeza et al., 2022). Although several high-risk clusters detected in Chile include the year 2009, they span extended periods that appear to extend beyond the immediate impact of that recessionary year. In Spain, the period 2009–2013 was predominantly one of economic recession (World Bank, 2026), but no high-risk suicide cluster occurred mainly during this period. This lack of association may be explained by the life stage of the studied age group. In Spain, young people typically achieve independence, marriage, or parenthood later—often in their thirties—compared to the other countries analyzed. Therefore, feelings of failure related to life milestones or pressures to meet family responsibilities—recognized suicide risk factors— may be less relevant in this younger cohort, potentially accounting for the observed results. Although Uruguay experienced an economic crisis very similar to Argentina’s, with consecutive years of recession from 1999 to 2002 (Antía, 2003; CEPAL, 2026), national youth suicide rates, for both women and men, tended to reach higher values in the later years of the study period. Thus, the spatiotemporal clusters detected in Uruguay reflect the geographic expression of these rising national trends. Across all four countries, there appears to be no clear interaction between economic crises and territorial socioeconomic inequalities that would result in a disproportionate increase in suicides during recessionary years in lower-socioeconomic-status areas.

Our results showed no statistically significant variations in population density between suicide clusters and the rest of the territory—except in Spain, where low-risk clusters were characterized by low population densities. Given limited access to mental health services, greater firearm availability, spatial isolation, depopulation, and other adverse conditions, high-risk clusters of youth suicide were expected in rural areas (Ghadipasha et al., 2024; Runkle et al., 2023). Although population density itself was not linked to elevated risk, the Patagonian region of Argentina—an area of low density and geographic isolation—did form a cluster of youth male suicides during 2002-2013. A similar pattern was observed in Chile, with a cluster emerging in its Patagonian region.

A common feature across all four countries is the absence of high-risk youth suicide clusters in their most populous urban centers. In Argentina, the Autonomous City of Buenos Aires registered a low-mortality cluster. In Chile, the province of Santiago—which contains the capital city—had no high-mortality cluster. Similarly, in Spain, the provinces of Madrid and Barcelona, and in Uruguay, the departments of Montevideo and Canelones (the latter part of the Montevideo metropolitan area), showed no elevated clusters. Unlike rural settings, major cities typically offer better mental healthcare access, higher overall well-being, and may serve as a refuge for young people facing stigma in rural or small-town environments.

The COVID-19 pandemic was anticipated to heighten suicide risk among youth by intensifying mental health strains linked to social isolation and economic instability—effects expected to be most severe in socially vulnerable populations (Bridge et al., 2023). However, our findings did not indicate that the COVID-19 pandemic triggered the emergence of high-risk suicide clusters. Instead, the pandemic period coincided with a decline in suicides within several existing high-mortality clusters: in Argentina (cluster 1 for women, clusters 6 and 7 for men) and in Chile (cluster 3 for women). During the pandemic, increased family communication, stronger cohesion, and enhanced perceived social support—protective factors—were reported (Reed et al., 2023). On the other hand, the clusters that emerged at the end of our study period arose several times before the COVID-19 pandemic, with onset ranging from 2010 (Argentina: women and men; Uruguay: men) to 2018 (Uruguay: women). This timing suggests that long-term social shifts, rather than the immediate effects of the pandemic, underlie the elevated suicide risk in these areas.

One of these long-term social changes associated with suicide is the weakening of social bonds. In the United States and Canada, long-term trends of increased youth suicide have been found to be linked to rising divorce rates and secularization (Stockard and O’Brien, 2002; Trovato, 1992). In our study, the percentage of unmarried people and the percentage of single-person households were both considered proxy variables for the level of social fragmentation in an area. In Argentina, a higher average percentage of unmarried people was observed in provinces located in clusters with a high risk of suicide among young women. In Spain, for women, this percentage was lower in high-risk suicide clusters compared to the rest of the country, while the percentage of single-person households was higher in high-risk clusters. These processes, associated with weakening community ties, unfold over extended periods and may follow a spatial-diffusion model, spreading from more economically developed areas toward more peripheral regions.

Although the increasing use of social media has been highlighted as a factor in the spread of more lethal suicide methods among young people, especially women (Ormiston et al., 2024), the spatial and temporal clustering observed in our study suggests that local geographic influences may also drive the diffusion of high-lethality means. However, empirical evidence regarding the role of social media in youth suicide incidence across these four countries remains absent.

This study has several limitations. First, although deaths by hanging, strangulation, and suffocation of undetermined intent were included in Argentina and Chile, problems of underreporting of suicides may persist. However, we have no evidence that this limitation— present across all four countries—introduces systematic geographical bias. Second, the spatial units of analysis contain internal sociodemographic heterogeneity in socioeconomic status, population density, and social fragmentation, which may mask more localized patterns. Third, the use of centroids in the spatiotemporal scan analysis could have influenced the size and duration of detected clusters; smaller geographical units might have revealed different spatial and temporal extents. Fourth, the selection of sociodemographic variables was constrained by their common availability across the four countries, so data on mental disorder prevalence, religious practices, and other relevant factors associated with youth suicide risk were not included. Such information is rarely available with both intra-national geographic detail and annual frequency in Latin American contexts.

Our results showed spatiotemporal variations in youth suicide across Argentina, Chile, Spain, and Uruguay. With the exception of Chile, recent national increases in youth suicide rates appear to be concentrated in specific sub-national regions, highlighting the importance of targeting resources to improve living conditions and access to mental healthcare for young people in these areas.

## Data Availability

The authors do not have permission to disseminate the research data.

## Statements and Declarations

### Competing Interests

the authors declare no conflict of interest.

### Funding

This research was funded by a post-doctoral fellowship awarded to Carlos M. Leveau by the *Fundación Carolina* (Spain) and the *Universidad Nacional de Lanús* (Argentina).

### Consent for publication

Not applicable.

### Ethics approval and consent to participate

Not applicable.

## Supplementary Material

**Table S1.**
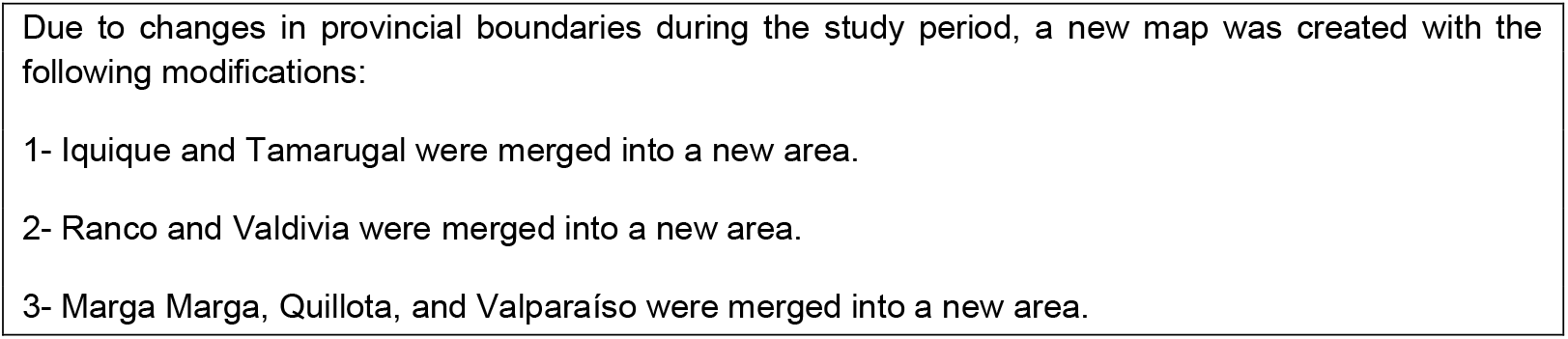
Changes in the composition of Chilean provinces.

**Table S2.** Suicides and deaths by hanging, strangulation and suffocation of undetermined intention* in a population aged 10-29 years. Argentina and Chile, 1997-2021.

| Year | Argentina |  |  |  | Chile |  |  |  |
| --- | --- | --- | --- | --- | --- | --- | --- | --- |
|  | Suicides (X60-X84) |  | Undetermined intent (Y20*) |  | Suicides (X60-X84) |  | Undetermined intent (Y20*) |  |
|  | Female | Male | Female | Male | Female | Male | Female | Male |
| 1997 | 158 | 388 | 21 | 89 | 63 | 259 | 21 | 128 |
| 1998 | 155 | 478 | 27 | 72 | 57 | 259 | 21 | 126 |
| 1999 | 141 | 519 | 14 | 76 | 54 | 254 | 22 | 134 |
| 2000 | 161 | 675 | 16 | 99 | 73 | 389 | 3 | 13 |
| 2001 | 228 | 738 | 13 | 33 | 101 | 427 | 0 | 1 |
| 2002 | 448 | 1648 | 7 | 34 | 82 | 416 | 0 | 0 |
| 2003 | 241 | 939 | 10 | 14 | 84 | 413 | 0 | 0 |
| 2004 | 248 | 964 | 3 | 20 | 105 | 449 | 0 | 0 |
| 2005 | 226 | 941 | 9 | 28 | 98 | 424 | 0 | 0 |
| 2006 | 261 | 976 | 10 | 34 | 103 | 419 | 0 | 0 |
| 2007 | 250 | 907 | 14 | 45 | 140 | 507 | 0 | 0 |
| 2008 | 264 | 954 | 21 | 127 | 167 | 548 | 0 | 0 |
| 2009 | 252 | 932 | 29 | 136 | 155 | 549 | 0 | 0 |
| 2010 | 216 | 1010 | 38 | 157 | 142 | 487 | 0 | 0 |
| 2011 | 259 | 1031 | 42 | 204 | 116 | 494 | 0 | 0 |
| 2012 | 280 | 1133 | 48 | 174 | 104 | 410 | 0 | 0 |
| 2013 | 257 | 1034 | 38 | 175 | 99 | 392 | 0 | 0 |
| 2014 | 308 | 1073 | 38 | 144 | 97 | 409 | 0 | 0 |
| 2015 | 242 | 955 | 44 | 178 | 111 | 392 | 0 | 0 |
| 2016 | 245 | 967 | 49 | 193 | 103 | 395 | 0 | 0 |
| 2017 | 291 | 1013 | 77 | 279 | 91 | 390 | 0 | 0 |
| 2018 | 290 | 1040 | 91 | 261 | 100 | 376 | 0 | 0 |
| 2019 | 264 | 1070 | 109 | 282 | 105 | 373 | 0 | 0 |
| 2020 | 212 | 928 | 97 | 277 | 94 | 310 | 0 | 0 |
| 2021 | 263 | 878 | 132 | 390 | 100 | 319 | 0 | 0 |

**Table S3.** Socio-demographic factors of the area used in the four countries, 1997-2021.

| Area factor | Variable | Sources |
| --- | --- | --- |
| Socioeconomic level: social and economic well-being of the population residing in an area. | Percentage of population with completed university or tertiary education. | Argentina: 2022 Census.<br>Chile: 2017 Census.<br>Spain: 2023 Annual Census.<br>Uruguay: 2011 Census. |
| Level of urbanization: understood as population concentration. | Population density: inhabitants per square kilometer. | Argentina: 2022 Census.<br>Chile: 2017 Census.<br>Spain: 2021 Annual Census.<br>Uruguay: 2011 Census. |
| Social fragmentation: a decrease in social ties within a population residing in an area. | Percentage of adult population that is single, divorced or widowed (unmarried or single population). | Argentina: 2010 Census.<br>Chile: 2002 Census.<br>Spain: 2020 Continuous Household Survey.<br>Uruguay: 2011 Census*. |
|  | Percentage of single-person households. | Argentina: 2022 Census.<br>Chile: 2017 Census.<br>Spain: 2020 Continuous Household Survey.<br>Uruguay: 2011 Census. |

\* The marital status variable was replaced by the variable "Spouse or partner in the home", because marital status showed anomalous results compared to the other countries: on average, only 5% of the population declared that they were married.

## Notes

### Competing Interest Statement

The authors have declared no competing interest.

### Funding Statement

This research was funded by a post-doctoral fellowship awarded to Carlos M. Leveau by the Fundacion Carolina (Spain) and the Universidad Nacional de Lanus (Argentina).

### Author Declarations

The datasets used in our study had been aggregated prior to use in our study.

